# Optimization of head and neck cancer peripheral blood mononuclear cell processing for robust downstream functional immune analysis

**DOI:** 10.64898/2026.04.29.26351856

**Authors:** Margaret Nelson, Katelyn Jansen, Farah Sagin, Maria Lehn, Hani Alrefai, Claire Girten, Joanna Khoury, Marina Rodriguez, Jacob Garner, Carolyn Schroeder, Melissa Meyer, Pravin Mishra, Dalia El-Gamal, Kelsey Dillehay McKillip, Trisha M Wise-Draper

## Abstract

The current “gold standard” for diagnosing and assessing treatment response is tumor biopsy; however, biopsies are not always feasible, safe or easily repeated during treatment. Utilization of peripheral blood mononuclear cells (PBMCs) as a surrogate for tumor biopsy allows for longitudinal sampling and is a safer, more readily available option. However, collection conditions, sample transfer time across multiple clinical sites, and PBMC processing conditions are external pre-analytical factors that must be understood and controlled to mitigate bias in downstream functional analyses. This study aims to systematically evaluate the pre-analytical variables affecting PBMC integrity and functional immune readouts as a prerequisite for downstream translational biomarker applications. Peripheral blood samples were collected from 80 treatment-naive patients with a diagnosis of head and neck squamous cell carcinoma. Blood was collected in cell preparation tubes (BD Vacutainer^®^ CPT™), potassium ethylenediaminetetraacetic acid (EDTA), or sodium heparin (SH) tubes and diluted 1:1 with sterile PBS or remained undiluted. PBMCs were processed and cryopreserved immediately or held for 8- and 24-hours before processing. PBMC viability was measured at cryopreservation and upon thawing. CD8+ T cells or natural killer (NK) cells derived from PBMCs were subjected to cytotoxicity assays using flow cytometry.

CPT™ tubes provided lower cell viability and yield at cryopreservation and upon thaw compared to EDTA and SH tubes while dilution had no effect on viability. NK cell cytotoxicity was similar between EDTA and SH tubes irrespective of dilution. However, diluted EDTA tubes resulted in lower T cell cytotoxicity after 24-hour hold. Viability and NK and T cell cytotoxicity were equivalent between cryopreserved PBMCs that were processed immediately or processed after 8- or 24-hour hold.

Here we report cryopreservation methods for reproducibility of viable cells that maintain functional immunological capacity even after significant delay in processing allowing flexibility and feasibility for collection from multiple clinical sites for deferred processing.

## Introduction

Understanding the mechanisms of anti-cancer treatment response is imperative in the design of novel cancer therapeutics. In addition to standard criteria such as stage and histology for choice of treatment, there is an increasing dependence on sophisticated patient sample analysis that allows detection of potential underlying drug targets and monitoring treatment response. Moreover, correlation of sample analysis with the clinical course allows for discovery of new biomarkers of treatment response and associated targetable pathways. Importantly, with the advent of immunotherapy, repeated sampling has become even more critical to understanding tumor and systemic immune response and to better predict immunotherapy response or mechanisms of resistance. Yet, unlike in hematological malignancies, a frequent limitation for solid tumors is that tissue biopsies are not always feasible, safe, permissible to the patient, or easily repeated during treatment. Therefore, there is an urgent need to establish reliable and valid alternatives to tissue biopsies.

While research into alternative ‘liquid biopsy’ methods such as circulating tumor cells (CTCs), cell free DNA (cfDNA), RNA and circulating exosomes are currently being investigated, major drawbacks of these methods include high cost, lack of standardization, and limitation in sampling for some tumor types ^1–3^. Therefore, shifting towards peripheral blood immune cells as a surrogate of tumor response has the potential to mitigate major drawbacks of other liquid biopsy methods. The use of patient peripheral blood mononuclear cells (PBMCs), which may be isolated from standard whole blood, is non-invasive, safe to collect, and more readily obtainable before, during, and after treatment.

Accordingly, the first step towards the utilization of PBMCs requires careful optimization, validation, and standardization of collection and processing procedures. As PBMCs are particularly prone to preanalytical bias and must be intact and viable for functional assays, differences in human biospecimen collection, time to processing, methods of PBMC processing and storage techniques represent the most significant sources of preanalytical variability. Previous work collected from healthy and diseased subjects has demonstrated that such preanalytical variables can confer detrimental effects on the viability of PBMCs and their suitability for functional assays ^4–6^. For instance, comparison between ethylenediaminetetraacetic acid (EDTA), acid-citrate-dextrose (ACD), and sodium heparin (SH) collection tubes revealed no significant differences in PBMC recovery or viability ^7^, however, blood collected in EDTA tubes tended to deteriorate faster with increasing time between collection and storage^8,9^. Yet, since EDTA chelates divalent metal ions, thereby inhibiting DNA and RNAses^10^, EDTA tubes have proven superior to SH or citrate tubes in preserving samples for downstream application of DNA and RNA analysis ^11,12^.

Additionally, processing blood within 8 hours of collection increases yield and viability of PBMCs while also minimizing contamination with granulocytes, which are potent suppressors of T-cell function ^5^ and are known to become activated in cancer^13^. However, immediate processing is often not a feasible approach as many facilities are not equipped for PBMC isolation and require the ability to ship whole blood to a central site necessitating optimized methods that allow delayed processing. Separation technique is another important consideration when preparing PBMCs for functional analyses. Isolation of PBMCs by density gradient centrifugation is considered the gold standard. However, the Ficoll separation method is known to be laborious and highly operator-dependent based on individual skill and technique. Comparison studies of commercial PBMC preparation tubes have demonstrated that these methods perform similar or better to the standard Ficoll separation method, with both SepMate™ and cell preparation tubes (BD Vacutainer^®^ CPT™) improving PBMC recovery, viability, and T-cell function ^14^. Based on previous findings, optimization of preanalytical variables is paramount in minimizing loss of functional outcomes and maximizing long-term viability for downstream analyses of immune function.

Therefore, the goal of this study was to elucidate the effects of preanalytical variables including blood collection tube type, effect of phosphate buffered saline (PBS) dilution, and time between collection and processing, on PBMC viability and downstream immune functionality. To this end, peripheral blood samples were collected from 80 patients with head and neck squamous cell carcinoma (HNSCC) and PBMCs were processed and compared across the tested variables. Overall, our findings highlight that blood collection tube has the greatest influence on PBMC viability and immune cell function. Additionally, the viability of PBMCs and cytotoxic capacity of NK and T cells are comparable between cryopreserved PBMCs that were immediately processed or held for 8- or 24-hours before processing. This apparent tolerance defines an operational window within which PBMC processing can be performed without compromising functional integrity. Our current study provides proof-of-principle that establishing and harmonizing standard procedures for processing PBMCs allows for reproducible collection of viable cells that maintain functional capacity upon storage and from which downstream functional assays can be performed. Moreover, the optimization and standardization of PBMC processing procedures herein allows for these methods to be adopted and utilized at different facilities, making it conducive for clinical trials and future standard PBMC functional testing for outcome and biomarker analysis.

## Materials and Methods

### Blood product procurement

Blood procurement involving human subjects was approved by the Institutional Review Board (IRB) of the University of Cincinnati (UCCI-UMB-14-01, IRB# 2015-2344 and General Specimen Protocol IRB# 2017-2137) and conducted in accordance with Good Clinical Practice guidelines and the Declaration of Helsinki. Written and informed consent was collected from enrolled patients with HNSCC prior to sample collection. Eligibility criteria included patients who were ≥ 18 years of age with a confirmed diagnosis of HNSCC with no prior chemotherapy or radiation treatment, no previous history of autoimmune disease, immunodeficiency disorders or immunosuppressive medications. Patient demographic information is summarized in **Supplemental Tables 1-4**.

### PBMC isolation

PBMCs from patients with HNSCC were processed by the University of Cincinnati Cancer Center Biospecimen Shared Resource at the University of Cincinnati (Cincinnati, OH). Whole blood was collected in either potassium EDTA tubes (Fisher Scientific, cat#02-657-32), SH tubes (Fisher Scientific, cat#02-689-6), or sodium heparin BD Vacutainer^®^ glass mononuclear CPT tubes (Fisher Scientific, cat#14-959-51D) and maintained at room temperature (22-30°C) prior to processing. Blood collected in BD Vacutainer^®^ CPT™ tubes were centrifuged at room temperature for 20 minutes at 1500xg within two hours of collection to separate the plasma and mononuclear cells. Additionally, to assess the effects of granulocyte contamination on viability, EDTA and SH blood were diluted 1:1 with sterile PBS + 2% heat-inactivated fetal bovine serum (hiFBS; Gibco, cat#A3382101) immediately following collection and prior to hold period. Blood was either processed immediately or at delayed timepoints (8 or 24 hours) to account for same-day and overnight processing delays, respectively. A schematic illustration of the processing workflow is provided in **Figure 1**.

**Figure 1:**
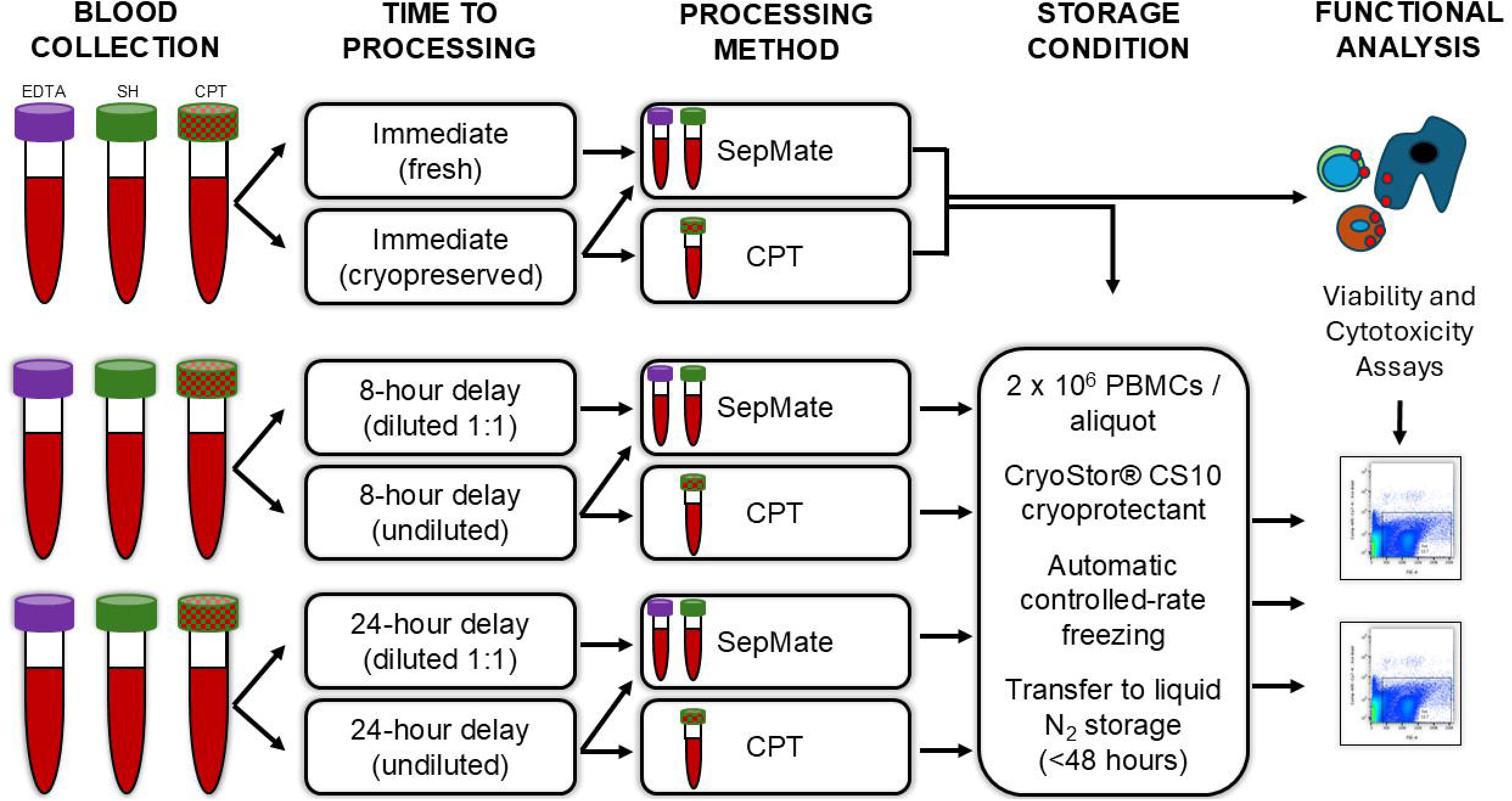
Schematic of workflow for blood collection and PBMC isolation. Blood was collected in three different tube types (EDTA, SH, or BD Vacutainer^®^ CPT™) and either processed immediately (within 30 minutes) or at delayed timepoints of 8- and 24-hours to account for both same-day and overnight processing delays, respectively. Whole blood was maintained at room temperature (22-30°C) prior to processing. Fresh PBMCs were used as a control to account for the effects of cryopreservation and processing delays on PBMC viability. The study measured cell viability and cytotoxicity assays across tube type, dilution, and processing delay.

The diluted and undiluted specimens were processed using SepMate™ tubes (StemCell Technologies, cat#85450) loaded with Ficoll-Paque™ PLUS media (Fisher Scientific, cat#45-001-749) and spun at 1,200xg at room temperature for 20 minutes. The supernatant containing plasma and PBMCs was collected and washed twice in a wash buffer consisting of PBS supplemented with 2% heat-inactivated fetal bovine with centrifugation at 300xg for 8 minutes. Cell counts and cell viability were assessed using the Denovix CellDrop Automated Cell Counter (DeNovix Inc., Wilmington, DE). After a final wash and spin, PBMCs were then resuspended in Cryostor^®^ CS10 cell freezing medium (StemCell Technologies, cat#07931) at a concentration of 2×10^6^ cells/mL and frozen at a rate of 1°C per minute until reaching −80°C using the StrexCell CytoSensei Controlled Rate Freezer (Strex USA, San Diego, CA). After storage for 48 hours at −80°C, PBMCs were transferred to vapor phase liquid nitrogen for long-term storage and provided for experimentation.

### Thawing and resting of PBMCs

Cryopreserved PBMCs were thawed and washed of freezing medium with RPMI-1640 (Fisher Sci, cat# MT10040CV) supplemented with 1% Penicillin/Streptomycin (Corning, cat#MT30002CI) and 10% hiFBS. PBMCs were counted and plated at a density of 1×10^6^ cells/mL in RPMI with 100U/mL of human recombinant IL-2 (rHu IL-2; Gibco, cat#200-02-250UG). PBMCs were rested overnight at 37°C in a humidified incubator with 5% CO_2_.

### Human cell lines

The human derived HNSCC cell line HN5 (gifted by Dr. Ben-Jonathon) were used as target cells for NK and T-cell cytotoxicity assays. HN5 cells were cultured and maintained in Dulbecco’s Modified Eagle Medium (DMEM, Corning, cat#MT10013CV) supplemented with 1% penicillin/streptomycin, 1% non-essential amino acids (Gibco, cat#11140050), 1mM sodium pyruvate (Corning, cat#25000CI), 2mM L-glutamine (Corning, cat#25005CI) and 10% hiFBS. Cells were maintained at 37°C with 5% CO_2_ during experimentation.

### NK and T cell cytotoxicity assays

HN5 cells were chosen based upon previous optimization of NK and T cell cytotoxicity assays (data not shown). HN5 cells were collected and resuspended in 0.5mM of 5 (6)-Carboxyfluorescein diacetate N-succinimidyl ester (CFDA-SE, Invitrogen, cat#V12883) diluted in PBS for 10 minutes at 37°C. HN5-CFSE stained cells were resuspended in complete DMEM with 200U/mL of rHu IL-2. Cells were plated in a 96-well plate at a density of 10,000 cells per well and were maintained at 37°C until NK or T cells were added.

NK cells were isolated from PBMCs per manufacturer’s instructions using the human NK cell isolation kit (Miltenyi, cat#130-092-657). Isolated NK cells were resuspended in DMEM with 200U/mL rHu IL-2 and co-cultured with HN5 cells at a density of 50,000 NK cells per well at a 5:1 effector: target (E:T) ratio for 4 hours at 37°C. After the 4-hour co-incubation cells were collected for flow cytometry.

T cells were isolated using the human T cell isolation kit (Miltenyi, cat#130-045-201). Isolated T cells were resuspended in DMEM with 200U/mL rHu IL-2 and co-cultured with HN5 cells at a density of 50,000 cells per well (5:1 E:T ratio). To enhance the interaction and recognition between allogenic T cells and target HN5 tumor cells, a CD44v6 Bispecific T-cell Engager (BiTE) was added to the co-culture. A CD19 B-cell specific BiTE and control media (medium used to develop the BiTEs but absent of BiTEs) were added to separate wells as additional controls. All BiTEs and control media were kindly provided by the Suzuki laboratory at Baylor University (Waco, TX) under a material transfer agreement. Details describing the generation of the BiTEs can be found in previous publications ^15,16^. Cells were co-cultured overnight before being collected.

Following the specific co-culture timeframe, target and effector cells were trypsinized, (0.25% trypsin, Corning cat#25-053-CI) collected in flow cytometry tubes (Corning Falcon, cat# 352054) and spun at 400xg for 5 minutes at 4°C. Cells were washed in flow cytometry buffer (PBS + 2% FBS, 1mM EDTA) and placed on ice. Cells, excluding unstained control and CFSE single stain control, were stained with 7AAD Viability Staining Solution (BioLegend, cat#420403) and immediately acquired on a LSR Fortessa (BD Biosciences, Franklin Lakes, NJ). Live target HN5 cells were considered as any cell that was CFSE+/7AAD- and dead target cells were identified as CFSE+/7AAD+. Basal cell death was calculated from CFSE-stained target cells without the addition of NK or T cells. Effector dependent cell death was calculated as the fold change of CFSE+/7AAD+ target cells over basal cell death. The gating strategy for the NK cell cytotoxicity assay (NKCA) and T cell cytotoxicity assays (TCCA) are shown in **Supplemental Figure S1**.

### Statistics

All data were analyzed using GraphPad Prism v10.6.1 (Boston, Massachusetts) and are represented as mean ± SEM. Two or more groups comprised of matching patients were analyzed using repeated measures two-way ANOVA. Paired Student’s t-test was used when comparing the matched patients at 0-, 8-, and 24-hour holds within the same assay. Significance was considered if *p*-value was less than 0.05 (*p*<0.05).

### Data Availability Statement

Patient-level deidentified datasets used during the current study are provided as supplemental tables. Additional information can be provided upon reasonable request to the corresponding author.

## Results

### PBMC viability is influenced by tube type and time after thaw

Since variables of blood collection (e.g., blood collection tube, time between collection and processing, etc.) can heavily influence the viability, recovery and immune functions of PBMCs, we first determined the optimal collection tubes for viability. Blood was collected in BD Vacutainer^®^ CPT™, SH or EDTA tubes and PBMC cell viability was compared between time at cryopreservation (immediately after PBMC processing and before controlled-rate freezing) and at thaw (when removed from liquid nitrogen, thawed, and washed of cryopreservation medium). Cell viability was equivalent between cryopreservation and at thaw when collected in SH or EDTA tubes but was significantly attenuated with CPT™ tubes (**Supplemental Figure S2 and Supplemental Table 5**). Based on these results, only SH and EDTA were used for subsequent experiments.

Next, PBMC viability was compared at three timepoints: at cryopreservation, at thaw, and after resting overnight. As expected, the highest cell viability was observed at cryopreservation and viability decreased approximately 10% upon thawing and after overnight incubation for both 8- and 24-hour holds across all tube types (**Figure 2A-B, Supplemental Tables 6-8**). Although not significant, there was a trend for lower viability at cryopreservation and after resting observed with SH tubes. After the 24-hour hold no significant difference was observed in cell viability between blood collection tubes at the three timepoints (**Figure 2B**).

**Figure 2:**
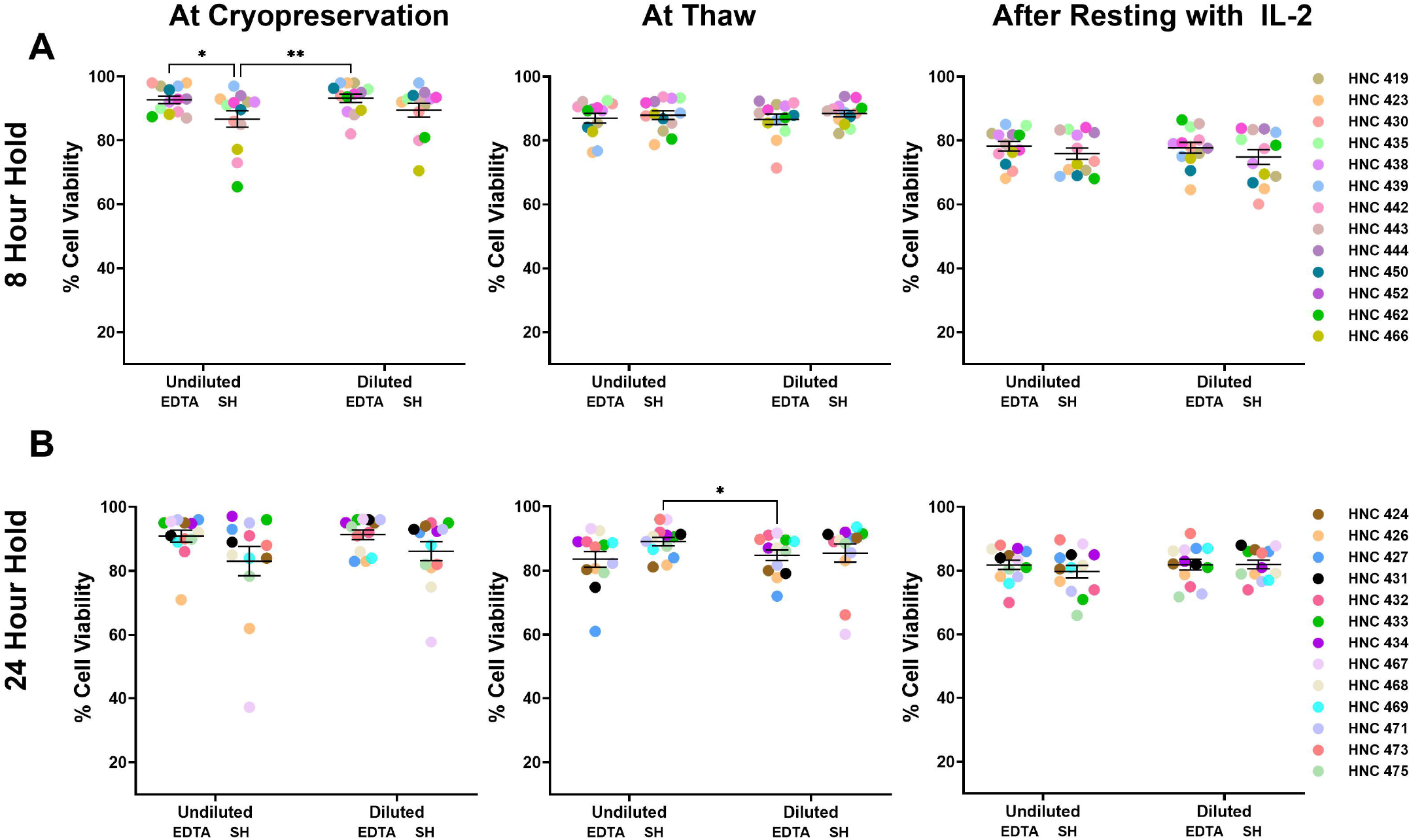
Comparison of PBMC viability across blood collection tube type and PBS dilution. Blood samples were collected and either held for (A) 8-hours or (B) 24-hours at room temperature (RT) before PBMC processing. PBMC cell viability was then measured at three experimental timepoints: at the point of cryopreservation, upon thawing from −80°C, and after overnight resting with rHu IL-2 (100U/μL) using Denovix CellDrop Automated Cell Counter. N=13 patients per time hold. Data were analyzed using a repeated measures two-way ANOVA. \**P*<0.05, \*\**p*<0.001.

### Dilution of whole blood samples did not improve viability

Since an increase in activation of CD11b+/CD15+ granulocytes leading to T-cell dysfunction has been observed in patients with cancer with advanced disease^13^, we tested whether diluting the samples with PBS would preserve PBMC viability. We first compared the effect of SH or EDTA blood collection tubes with or without PBS dilution after 8- or 24-hour hold at room temperature. We found that after the 8-hour hold, undiluted SH tubes showed a lower viability only at cryopreservation, but viability was comparable between the diluted and undiluted tubes at thaw and after resting. For the 24-hour hold, cell viability was not affected by dilution or tube type (EDTA and SH) at any of the timepoints (**Figure 2A-B, Supplemental Tables 6-7**). Although inter-patient variability was observed, consistent trends across matched samples indicate that the effects of processing conditions were not driven by individual patient differences.

### NK and T cell cytotoxicity is not improved by SH tubes or dilution

After establishing how blood collection tubes can influence cell viability, we next investigated how these variables impact PBMC immune cell function. NK and T-cells were isolated from PBMCs, and cytotoxicity was determined using flow cytometry. NK cell cytotoxicity was not altered by blood collection tube (EDTA and SH) or PBS dilution, and cytotoxicity was retained between the 8- and 24-hour holds (**Figure 3A**). For T cell cytotoxicity, no statistical difference was observed between tube type and dilution after the 8-hour hold, yet after the 24-hour hold, a significant decrease in T cell cytotoxicity was seen from EDTA tubes diluted with PBS compared to undiluted EDTA tubes (**Figure 3B**). Based on these initial findings which showed no additional benefit in viability or T cell function by dilution or using SH tubes, combined with previous studies showing that EDTA preserves RNA and DNA better than other collection tube types^10,12^, undiluted EDTA tubes were selected for the remaining experiments.

**Figure 3:**
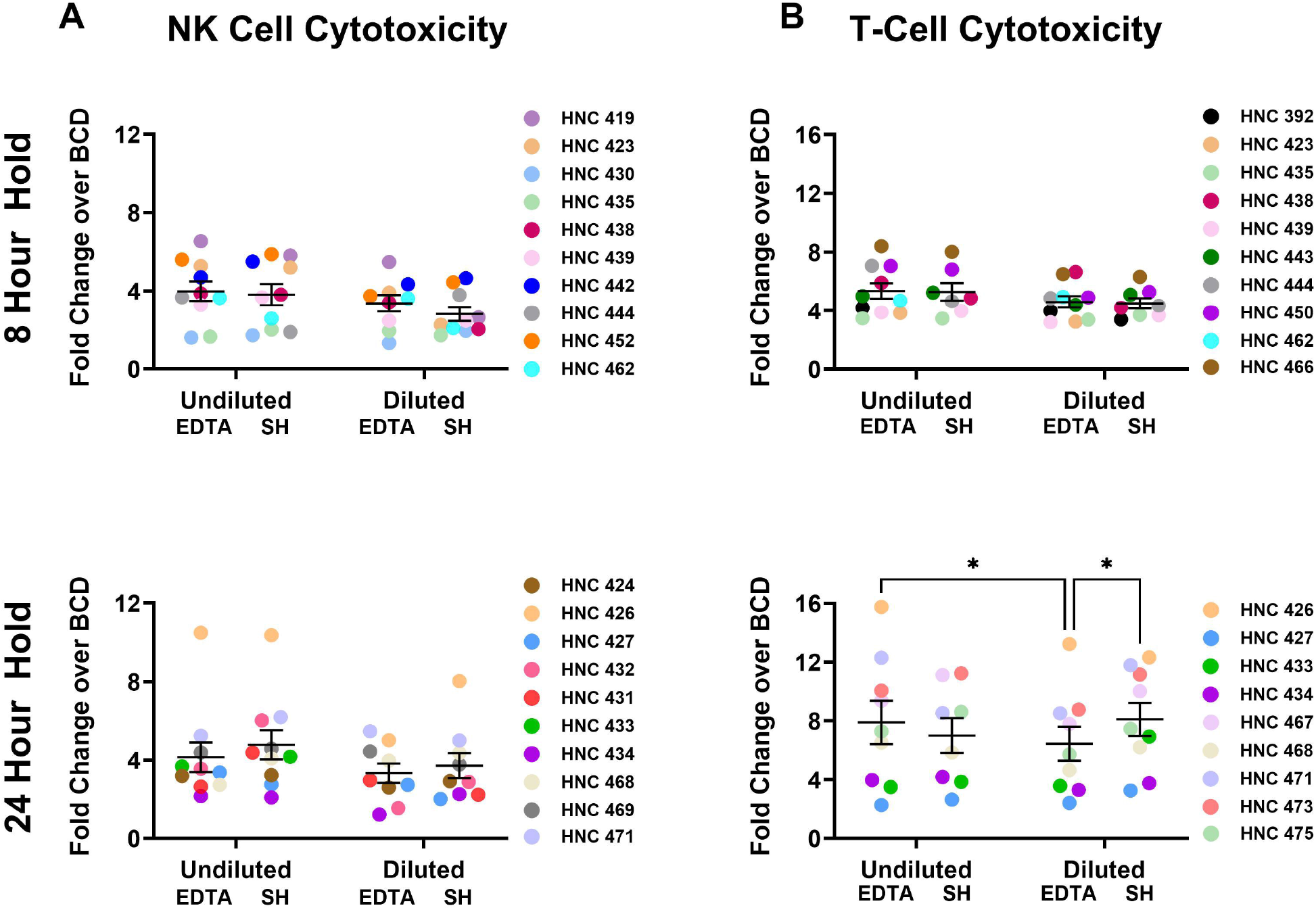
NK and T-cell cytotoxicity are retained from undiluted EDTA collection tubes. (A) NK cells or (B) T cells were isolated from PBMCs processed after 8- or 24-hours. NK cells were co-cultured with CFSE-stained HN5 tumor cells. NK-dependent tumor cell death was measured by calculating the fold change over the basal cell death (BCD) of HN5 cells without the addition of NK cells. T cells were co-cultured with CFSE-stained HN5 tumor cells and anti-CD44 BiTE, anti-CD19 BiTE, or control media (no BiTE). T cell dependent tumor cell death was measured as the fold change of the CD44-BiTE over basal cell death. N=10 for NK cytotoxicity assays and n=9 for T cell cytotoxicity assays. Data were analyzed using a repeated measures two-way ANOVA. \**P*<0.05.

### NK and T cell cytotoxicity is preserved with the 24-hour hold processing conditions

To account for the effects of cryopreservation and processing delays on immune function of PBMCs, we next sought to compare these findings to freshly isolated PBMCs. For these experiments, PBMCs from the same patient were either processed immediately after collection and analyzed by cytotoxicity assay (fresh) or processed and cryopreserved immediately (0 hour) or processed after delay (8- and 24-hour hold). The ratio of cell death across controls (Ctl and CD19 BiTE) compared to CD44 BiTe (T cell dependent death) were equivalent across the 0-, 8- and 24-hour time points (**Figure 4A, Supplemental Figure S3**). In order to confirm that NK cell cytotoxicity was also preserved with delay in processing, a NKCA was also performed to compare fresh cryopreserved PBMCs to PBMCs processed after a 24-hour hold. The NKCA showed similar findings as the T cell cytotoxicity assay, confirming that cryopreserved PBMCs with 0- or 24-hour hold have comparable immune effector functions (**Figure 4B**).

**Figure 4:**
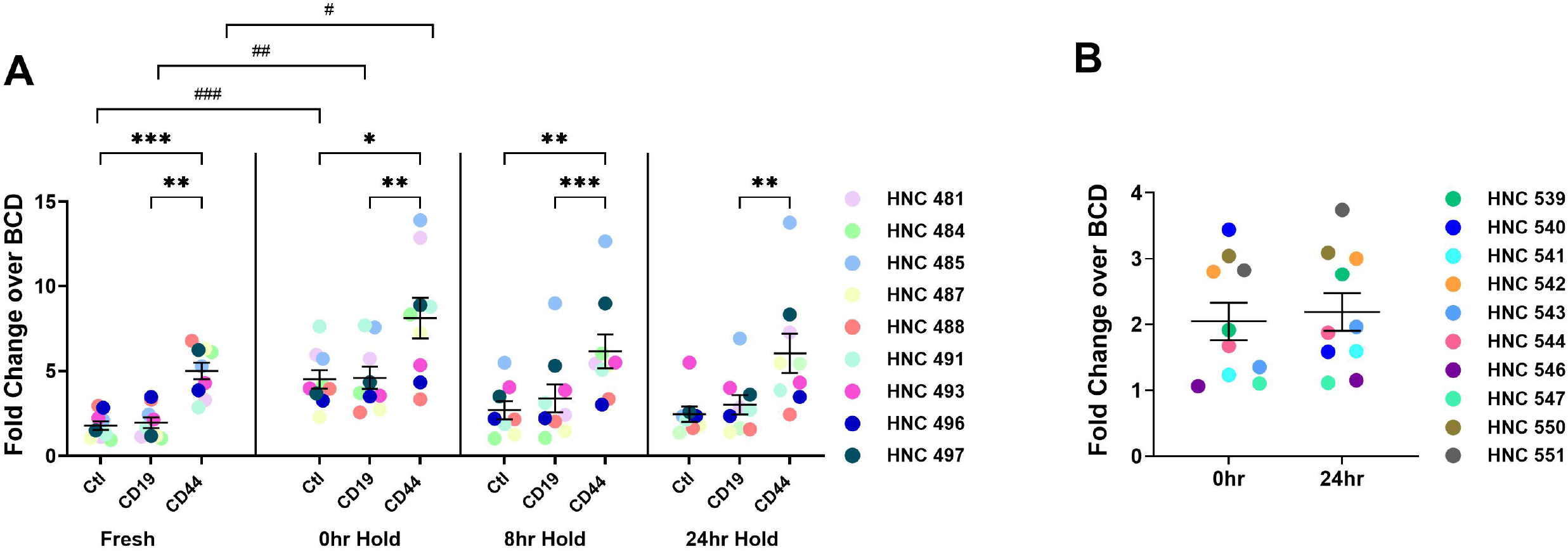
NK and T cell cytotoxicity are comparable between immediate processing, and 24-hour hold PBMC processing. (A) T cells were isolated from PBMCs that were either processed immediately (fresh) or thawed after 0-, 8-, or 24-hour hold prior to PBMC isolation and cryopreservation. T cells were co-cultured with CFSE-stained HN5 tumor cells and anti-CD44 BiTE, anti-CD19 BiTE, or control media (no BiTE). T cell dependent tumor cell death was measured as the fold change of the CD44-BiTE over basal cell death. (B) NK cells were isolated from PBMCs that were cryopreserved immediately or after a 24-hour hold. NK cells were co-cultured with CFSE-stained HN5 tumor cells. NK-dependent tumor cell death was measured by calculating the fold change over the basal cell death (BCD) of HN5 cells without the addition of NK cells. N=9 for T cell cytotoxicity assays and n=10 for NK cell cytotoxicity assays. 0-, 8-, and 24-hour hold data were analyzed using a repeated measures two-way ANOVA with Tukey’s multiple comparison test. \**P*<0.05, \*\**p*<0.01, \*\*\**p*<0.001. Comparison of Ctl, CD19 or CD44 from fresh and 0hr hold were compared and analyzed using unpaired Student’s t-test. #*p*<0.05, ##*p*<0.01, ###*p*<0.001. 0- and 24-hour hold data from NK cytotoxicity assay was compared using paired Student’s t-test.

## Discussion

The aim of this study was to test and identify variables in blood collection and processing that would both preserve viability and functionality while ensuring the methods were feasible and could be easily replicated at other sites especially after delay to allow for shipment.

We first compared PBMC viability from BD Vacutainer^®^ CPT™, EDTA and SH blood collection tubes. Results showed that PBMC yield and viability were significantly reduced when blood was collected from CPT™ tubes after 8- and 24-hour processing delay. Previous studies have also observed similar reduction in cell yield with CPT™ tubes ^17^. Although CPT™ tubes contain the density gradient allowing for cell separation in the same tube, continuous exposure to such gradient may inversely affect cell viability. Conversely, in our study EDTA tubes maintained the greatest PBMC yield and viability at cryopreservation, upon thaw and after resting compared to SH tubes. Additionally, compared to SH tubes, EDTA tubes have proven suitable for retaining quality RNA and DNA for downstream applications^10,12^, while heparin has been found to disrupt PCR reactions by inhibiting Taq DNA ^18,19^. Given that our current protocol using undiluted whole blood includes the option of an additional spin to preserve plasma, EDTA tubes appear to be the most beneficial in both the functionality and viability of PBMCs as well as downstream genome and transcriptome processing. Several studies have also previously shown that extending time between collection and processing leads to contamination of granulocytes, which can suppress T-cell function, specifically within cancer ^5,13^ and can be overcome with PBS dilution. Yet, we observed no additional benefit in viability or T-cell functionality when blood was diluted with PBS prior to processing. Taken together, these findings suggest EDTA tubes perform better at maintaining PBMC cell viability and immune effector function with blood samples from patients with HNSCC.

Within this study we also tested the effect of delayed PBMC processing (8- and 24-hour hold) on immune cell functionality. These timepoints were chosen to represent the timing of same-day shipping (8 hours) or overnight shipping (24 hours) if samples were collected and sent to a central site for analysis. We chose these timepoints for analysis since, in the context of multicenter clinical trials and analytical validation of immune-based biomarkers, immediate and onsite processing is not always feasible and poses a significant logistical challenge. Our NK and T-cell functionality showed similar cytotoxicity capacities between cryopreserved PBMCs that were either immediately processed or held for 8- and 24-hours prior to processing. Although other studies have observed that granulocytes can become activated with longer blood storage time^5^, their density may decrease, and they may thus contaminate the enriched PBMC fraction, our finding that the viability and functionality was unchanged suggests that there was minimal sample contamination with granulocytes even after 24-hour hold. Within our study we also compared NK and T-cell cytotoxicity between freshly processed PBMCs (fresh) and PBMCs cryopreserved after processing (0-hour hold) or after 8- or 24-hour hold. Interestingly, T cells isolated from fresh PBMCs showed significantly lower cytotoxicity within the media (Ctl) and CD19 BiTE controls (**Figure 4A**), compared to immediately cryopreserved PBMCs (0-hour hold). For the NK cytotoxicity, similar to T cells, no difference was observed between the 0-hour and 24-hour hold. Previous studies have shown that thawing PBMCs followed by overnight resting significantly improves the immunogenicity of CD8+ T cells in response to various stimuli ^20,21^. Additionally, compared to fresh PBMCs, previously cryopreserved PBMCs have been shown to produce greater levels of cytotoxic cytokines including TNF-α and IFN-y ^22^ possibly contributing to increased cytotoxicity as observed in our study. Our findings suggest that there is at least a 24-hour window in which PBMC functionality and viability are preserved, which could be beneficial in multisite clinical trials and once approved as standard of care for community sites in which resources to process and analyze blood samples are limited. Future studies to test longer processing delays are needed and can provide a maximal timeframe for blood processing.

One important aspect of sample quality for analysis is the ability to retrieve healthy PBMCs after cryopreservation. We had previously compared the Mr. Frosty™ freezing container technique to the slow rate freezer and found that the retrieval was overall similar but improved with the slow rate freezer and as well as more efficient (data not shown). In our current study, the cryopreservation protocol adhered to stringent methods which utilized CryoStor^®^ CS10, a commercially available cryopreservative^®^. In addition, PBMCs aliquots first underwent controlled rate freezing after which they were held at −80°C for 48 hours and transferred to vapor phase liquid nitrogen storage until samples were ready for downstream use and functional analysis. Although these parameters were not tested in this study, we believe these to be optimal cryopreservation conditions as both viability and functionality were preserved across the different delays in blood sample processing.

There are some limitations to our study. We only used samples from patients with HNSCC and therefore, our results need to be verified across multiple tumor types. In addition, it is possible that by increasing patient numbers there may be significant differences between diluted and non-diluted samples as well as in NK and T cell cytotoxicity readouts between timepoints. Finally, other modifications during processing also could affect outcomes including storage media, various freezing techniques, etc. It will be important to expand on these limitations as well as lengthen processing delays to ensure applicability of our methods broadly to the community.

In summary, this study highlights the importance and difficulties of standardizing pre-analytical variables in order to optimize blood collection and PBMC isolation. Findings from our study can contribute to establishing PBMCs as a valid and safer option of ‘liquid biopsy’ while also providing a harmonized methodology of blood collection and processing that preserves viability and functionality of PBMCs.

## Acknowledgments

This work was supported by the National Cancer Institute (U01 CA267985-01). This publication’s contents are the sole responsibility of the authors and do not necessarily represent the official view of the NIH. The authors would also like to acknowledge the University of Cincinnati Clinical Trials Office translational clinical research coordinators for their assistance in consenting and enrolling patients.

## Notes

**COI/Disclosures:** TWD relationships are not relevant to this manuscript. The remaining authors declare that the research was conducted in the absence of any commercial or financial relationships that could be construed as a potential conflict of interest.

### Competing Interest Statement

The authors have declared no competing interest.

### Funding Statement

This work was supported by the National Cancer Institute (U01 CA267985-01). This publication contents are the sole responsibility of the authors and do not necessarily represent the official view of the NIH.

### Author Declarations

Blood procurement involving human subjects was approved by the Institutional Review Board (IRB) of the University of Cincinnati (UCCI-UMB-14-01, IRB# 2015-2344 and General Specimen Protocol IRB# 2017-2137) and conducted in accordance with Good Clinical Practice guidelines and the Declaration of Helsinki. Written and informed consent was collected from enrolled patients with HNSCC prior to sample collection.

### Summary of Updates

Revision is submitted to fix the supplemental figures that were inadvertently left off. Figures are now embedded in supplement.

