## Supplemental Tables and Figures for "Optimization of head and neck cancer peripheral blood mononuclear cell processing for robust downstream functional immune analysis"

**Supplemental Table 1: Patient demographics of PBMCs processed after 8-hour hold for use in the NK and T-cell cytotoxicity assays.**

| Patient | Age Range | Sex | Race | Smoking History (>10pk per year) | Alcohol History (>5 drinks per week) | ECOG Performance Status | Clinical Stage | Primary Disease Site | P16 Status |
|---|---|---|---|---|---|---|---|---|---|
| HNC 392 | 60-64 | Female | White | Yes | No | 1 | T3N0M0 | Larynx | Neg |
| HNC 419 | 65-69 | Male | White | No | No | 0 | T4aN2bM0 | Oral Cavity | Unk |
| HNC 423 | 60-64 | Male | White | No | Yes | 0 | T4aN1M0 | Oral Cavity | Unk |
| HNC 430 | 75-79 | Male | Other | No | No | 0 | T2N0M0 | Oral Cavity | Neg |
| HNC 435 | 75-79 | Female | White | No | No | 1 | T1N0M0 | Oral Cavity | Unk |
| HNC 438 | 65-69 | Male | White | No | No | 1 | T2N0M0 | Oral Cavity | Neg |
| HNC 439 | 80-84 | Male | White | No | No | 2 | T4aN0M0 | Larynx | Unk |
| HNC 442 | 65-69 | Male | White | No | Yes | 0 | TxN1M0 | Oropharynx | Pos |
| HNC 443 | 55-59 | Female | White | Yes | No | 1 | T2N0M0 | Oral Cavity | Unk |
| HNC 444 | 70-74 | Male | White | Yes | Yes | 1 | T2N0M0 | Oral Cavity | Unk |
| HNC 450 | 65-69 | Male | White | Yes | Yes | 0 | TxN1M0 | Oral Cavity | Unk |
| HNC 452 | 70-74 | Male | White | Yes | Yes | 0 | T2N0M0 | Oral Cavity | Unk |
| HNC 462 | 60-64 | Female | White | Yes | Yes | 0 | T1N0M0 | Oral Cavity | Unk |
| HNC 466 | 55-59 | Male | White | No | No | 0 | T2N0M0 | Oral Cavity | Unk |

| Supplemental Table 2: Patient demographics of PBMCs processed after 8-hour hold for NK and T-cell cytotoxicity assays. | | | | | | | | | |
|---|---|---|---|---|---|---|---|---|---|
| Patient | Age | Sex | Race | Smoking History (>10pk per year) | Alcohol History (>5 drinks per week) | ECOG Performance Status | Clinical Stage | Primary Disease Site | P16 Status |
| HNC 424 | 85-89 | Male | White | No | No | 1 | T2N1Mx | Oral Cavity | Unk |
| HNC 426 | 45-49 | Male | Black | No | Yes | 1 | T1N0M0 | Oral Cavity | Neg |
| HNC 427 | 50-54 | Female | White | No | No | 1 | T4aN2bM0 | Oral Cavity | Unk |
| HNC 431 | 45-49 | Male | White | No | No | 0 | T4N2M0 | Oral Cavity | Unk |
| HNC 432 | 80-84 | Male | White | No | No | 1 | TxN0M0 | Salivary Duct Carcinoma | Unk |
| HNC 433 | 40-44 | Male | White | Yes | No | 0 | T1N0M0 | Oral Cavity | Unk |
| HNC 434 | 70-74 | Female | White | Yes | Yes | 1 | T1N0M0 | Oral Cavity | Unk |
| HNC 467 | 70-74 | Female | White | Yes | No | 0 | T2N0M0 | Oral Cavity | Unk |
| HNC 468 | 55-59 | Male | White | Yes | No | 1 | T2N0M0 | Oral Cavity | Neg |
| HNC 469 | 65-69 | Male | White | No | No | 1 | T3N1M0 | Larynx | Neg |
| HNC 471 | 50-54 | Male | White | No | No | 1 | T2N0M0 | Oropharynx | Pos |
| HNC 473 | 55-59 | Male | White | No | No | 0 | T2N1M0 | Oropharynx | Pos |
| HNC 475 | 70-74 | Male | White | Yes | No | 0 | T1N0M0 | Nasal | Unk |

**Supplemental Table 3: Patient demographics of PBMCs processed either immediately (fresh) or processed and cryopreserved after 0-, 8- or 24-hour hold for use in T-cell cytotoxicity assays.**

| Patient | Age | Sex | Race | Smoking History (>10pk per year) | Alcohol History (>5 drinks per week) | ECOG Performance Status | Clinical Stage | Primary Disease Site | P16 Status |
|---|---|---|---|---|---|---|---|---|---|
| HNC 481 | 70-74 | Male | White | No | No | 0 | T2N1M0 | Oropharynx | Pos |
| HNC 484 | 60-64 | Male | White | No | Yes | 0 | T2N1M0 | Oral Cavity | Unk |
| HNC 485 | 90-94 | Female | White | No | No | 1 | T3N0M0 | Other-Sinonasal | Unk |
| HNC 487 | 85-89 | Female | White | No | No | 0 | T4N0M0 | Salivary Clear Cell Carcinoma | Neg |
| HNC 488 | 65-69 | Male | White | No | Yes | 1 | T4aN2aM0 | Larynx | Unk |
| HNC 489 | 50-54 | Male | White | Yes | Yes | 0 | T3N2bM0 | Oral Cavity | Unk |
| HNC 491 | 60-64 | Female | White | Yes | No | 1 | T2N0M0 | Oral Cavity | Unk |
| HNC 493 | 60-64 | Female | White | Yes | No | 1 | T2N0M0 | Oral Cavity | Unk |
| HNC 496 | 55-59 | Male | White | No | Yes | 0 | T2N1M0 | Oropharynx | Pos |

**Supplemental Table 4: Patient demographics of PBMCs processed and cryopreserved after 0- or 24-hour hold for use in the NK cytotoxicity assays.**

| Patient | Age | Sex | Race | Smoking History (>10pk per year) | Alcohol History (>5 drinks per week) | ECOG Performance Status | Clinical Stage | Primary Disease Site | P16 Status |
|---|---|---|---|---|---|---|---|---|---|
| HNC 539 | 25-30 | Male | White | No | No | 0 | T1N1M0 | Oropharynx | Pos |
| HNC 540 | 60-64 | Male | White | Yes | No | 0 | T3N2bcM0 | Larynx | Unk |
| HNC 541 | 70-74 | Male | White | Yes | Yes | 2 | T4bN3M0 | Larynx | Neg |
| HNC 542 | 75-79 | Male | White | Yes | No | 0 | T2N0M0 | Oral Cavity | Unk |
| HNC 543 | 55-59 | Male | White | Yes | No | 0 | T2N1cM0 | Oropharynx | Pos |
| HNC 544 | 55-59 | Male | White | No | No | 0 | T2N1M0 | Oral Cavity | Unk |
| HNC 546 | 60-64 | Male | White | No | No | 0 | T1N0M0 | Oropharynx | Pos |
| HNC 547 | 55-59 | Male | White | No | Yes | 0 | T2N1M0 | Oropharynx | Pos |
| HNC 550 | 60-64 | Female | White | Yes | No | 0 | T2N0M0 | Oral Cavity | Unk |
| HNC 551 | 65-69 | Male | White | No | No | 0 | T1N0M0 | Oral Cavity | Unk |

| Supplemental Table 5: Comparison of PBMC viability upon cryopreservation across collection tube types after 24-hour hold. | | |
|---|---|---|
| **Tube Condition** | **At cryopreservation (% live: mean ± SEM)** | **At thaw (% live: mean ± SEM)** |
| BD Vacutainer® CPT™ | 78.3 ± 7.42 | 48.7 ± 4.50 |
| EDTA | 93.3 ± 0.80 | 80.0 ± 1.91 |
| SH | 84.7 ± 3.30 | 84.0 ± 3.11 |

| Supplemental Table 6: Comparison of PBMC viability using EDTA or SH tubes +/- dilution after 8-hour hold. | | | |
|---|---|---|---|
| **Tube Condition** | **At Cryopreservation (% live: mean ± SEM)** | **At Thaw (% live: mean ± SEM)** | **After Resting (% live: mean ± SEM)** |
| EDTA - Undiluted | 92.7% ± 1.15 | 87.9% ± 1.75 | 77.9% ± 1.89 |
| SH - Undiluted | 86.7% ± 2.58 | 89.3% ± 0.94 | 75.5% ± 2.40 |
| EDTA - Diluted | 93.2% ± 1.33 | 87.3% ± 2.18 | 78.6% ± 1.67 |
| SH - Diluted | 89.5% ± 2.13 | 88.4% ± 1.44 | 73.4% ± 2.89 |

| Supplemental Table 7: Comparison of PBMC viability using EDTA or SH tubes +/- dilution after 24-hour hold. | | | |
|---|---|---|---|
| **Tube Condition** | **At Cryopreservation (% live: mean ± SEM)** | **At Thaw (% live: mean ± SEM)** | **After Resting (% live: mean ± SEM)** |
| EDTA - Undiluted | 91.2% ± 1.51 | 85.2% ± 1.73 | 82.4% ± 1.71 |
| SH - Undiluted | 86.2% ± 2.98 | 86.4% ± 3.00 | 82.2% ± 1.54 |
| EDTA - Diluted | 91.0% ± 1.78 | 84.1% ± 2.39 | 82.0% ± 1.47 |
| SH - Diluted | 83.0% ± 4.57 | 90.5% ± 1.24 | 79.6% ± 1.99 |

**Supplemental Table 8: Comparison of PBMC viability at Fresh, 0-, 8-, and 24-hour hold.**

| Time Condition | At Cryopreservation (% live: mean ± SEM) | At Thaw (% live: mean ± SEM) | After Resting (% live: mean ± SEM) |
|---|---|---|---|
| 0 hr-hold | 94.0% ± 1.61 | 84.4% ± 2.11 | 72.3% ± 4.82 |
| 8 hr-hold | 91.1% ± 1.90 | 83.9% ± 2.47 | 78.7% ± 4.65 |
| 24 hr-hold | 95.9% ± 1.06 | 81.0% ± 1.85 | 72.0% ± 2.65 |

**Supplemental Figures**

**Supplemental Figure S1. Gating strategy for NK and T-cell cytotoxicity assays.**

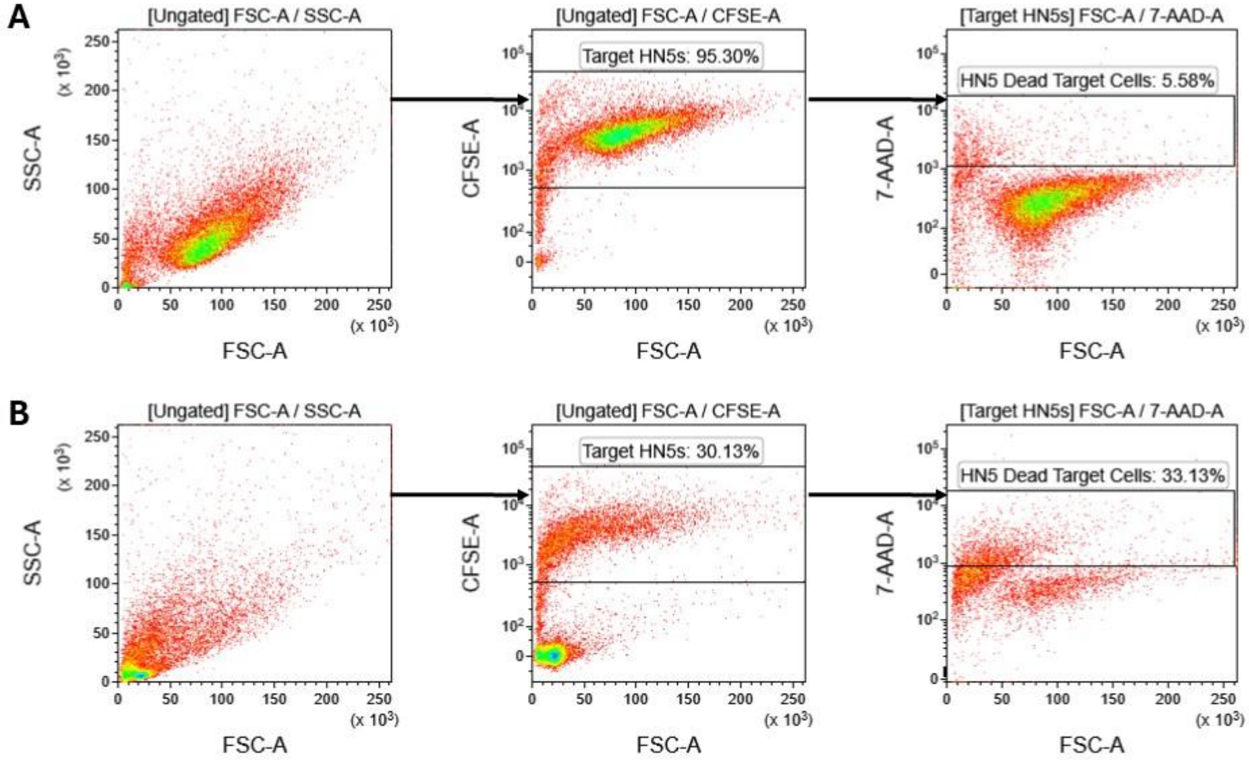


**Supplemental Figure S1: Gating strategy for NK and T-cell cytotoxicity assays.** Isolated NK or T-cells were co-cultured with CFSE-stained HN5 target cells at an effector: target ratio of 5:1. Cells were collected, stained with 7-AAD viability dye, and ran on a flow cytometer. Target HN5 cells were identified as CFSE+. Dead HN5 target cells were identified as CFSE+/7AAD+. (A) Gating strategy for target HN5 basal cell death (in the absence of effector cells). (B) Gating strategy of effector cell dependent cell death. Effector cell cytotoxic capacity was calculated as fold change of basal tumor cell death over effector cell dependent cell death. N=5 PBMCs collected in each tube type. (A) Data were analyzed using a repeated measures one-way ANOVA. (B) Data were analyzed using paired Student's t-test $*p<0.05$.

**Supplemental Figure S2. Comparison of PBMC yield and viability across tube types.**

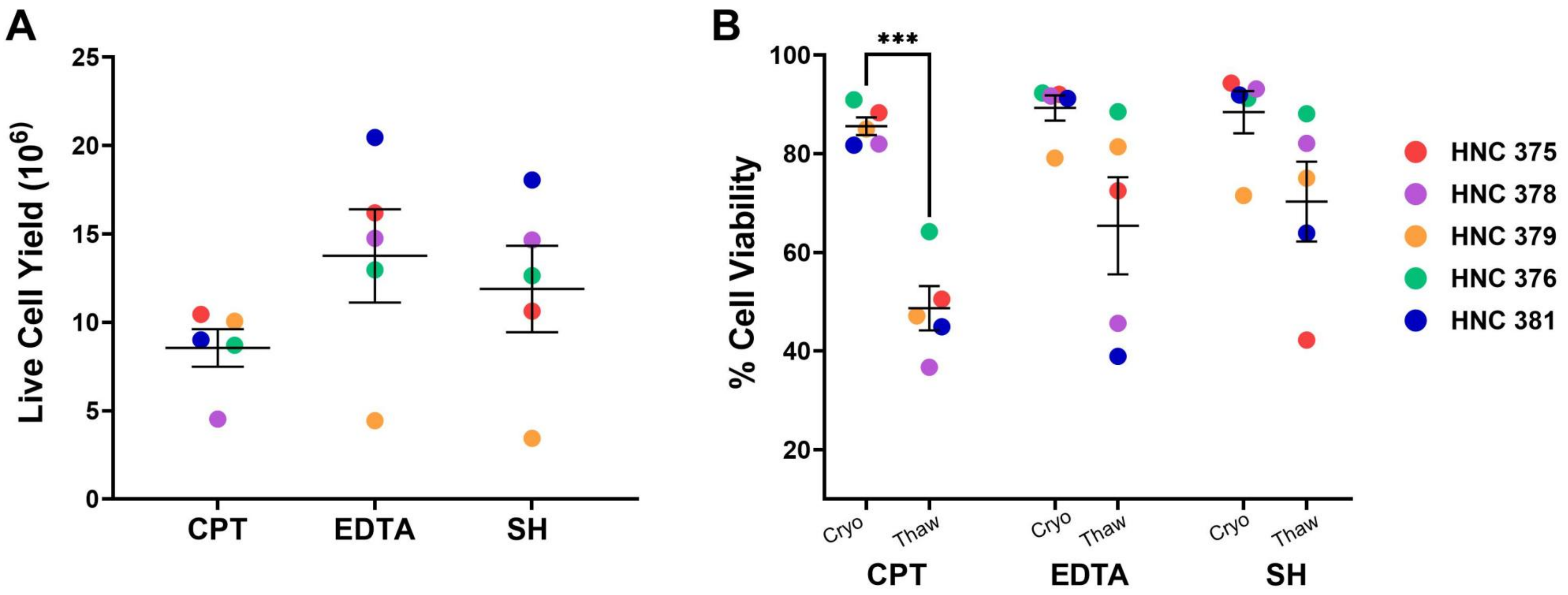


**Supplemental Figure S2: Comparison of PBMC yield and viability across tube types.** (A) PBMCs were processed after 24-hour hold and live cell yield was measured. (B) PBMC viability was measured upon cryopreservation and after thawing and compared at the two timepoints.

### Supplmental Figure S3: Comparison of T-cell cytotoxicity across PBMC processing delays.

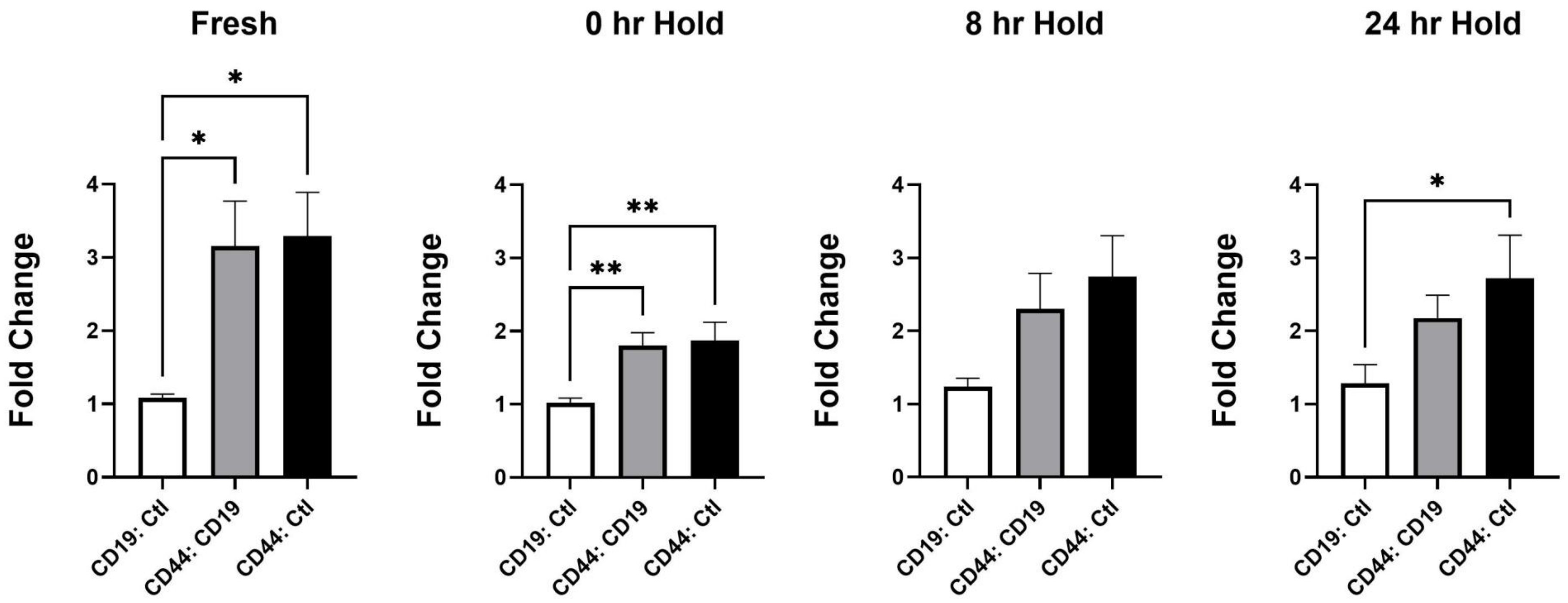


**Supplemental Figure S3: Comparison of T-cell cytotoxicity across PBMC processing delays.** Blood was collected and PBMCs were either processed and cytotoxicity was analyzed immediately (fresh), processed and cryopreserved immediately (0 hr-hold), or processed after 8- or 24-hour hold and cryopreserved. For the three cryopreserved timepoints, PBMCs were thawed simultaneously from each timepoint, and a T-cell cytotoxicity assay was performed. T-cell cytotoxicity was calculated as fold change of control, CD19 BiTE or CD44 BiTE over basal tumor cell death and presented as comparison of control or BiTEs. Data were analyzed using a repeated measures ANOVA with Tukey's multiple comparison test. *$P<0.05$, **$p<0.01$**.**
